# Relationship of sleep homeostasis to seizures and cognition in children with focal epilepsy

**DOI:** 10.1101/2020.11.05.20226514

**Authors:** Maria H Eriksson, Torsten Baldeweg, Ronit Pressler, Stewart G Boyd, Reto Huber, J Helen Cross, Bigna K Bölsterli, Samantha YS Chan

**Affiliations:** Developmental Neurosciences Programme, UCL Great Ormond Street Institute of Child Health, London, UK; Neuropsychology, Great Ormond Street Hospital NHS Trust, London, UK; Neurophysiology, Great Ormond Street Hospital NHS Trust, London, UK; Neurology, Great Ormond Street Hospital NHS Trust, London, UK; Pediatric Sleep Disorders Center, University Children’s Hospital Zurich, Zurich, Switzerland; Department of Neurology, University Children’s Hospital Zurich, Zurich, Switzerland; Children’s Research Center, University Children’s Hospital Zurich, Zurich, Switzerland; Child Development Center, University Children’s Hospital Zurich, Zurich, Switzerland; Department of Child and Adolescent Psychiatry and Psychotherapy, Psychiatric Hospital, University of Zurich, Zurich, Switzerland; Young Epilepsy, Lingfield, UK

**Author notes:** Corresponding author’s contact information: Ms Maria Eriksson, Developmental Neurosciences Programme, UCL Great Ormond Street Institute of Child Health, 30 Guilford Street, London, UK, WC1N 1EH. joint last authors (Bigna K Bölsterli, Samantha YS Chan).

**Keywords:** Epilepsy, sleep, children, EEG, slow wave activity, cognition, epileptiform discharges

## Abstract

**Objective:** Sleep disruption and cognitive impairment are important co-morbidities in childhood epilepsy, yet a mechanistic link has not been substantiated. Slow wave activity during sleep and its homeostatic decrease across the night is associated with synaptic renormalisation, and shows maturational changes over the course of childhood. Here, we aimed to investigate the effect of epilepsy on sleep homeostasis in the developing brain.

**Methods:** We examined the relationship of sleep homeostasis as reflected in slow wave activity to seizures, cognition and behaviour, comparing 22 children (aged 6 to 16 years) with focal epilepsy to 21 age-matched healthy controls. Participants underwent overnight sleep EEG and IQ testing and performed memory consolidation tasks. Their parents completed standard behavioural questionnaires.

**Results:** Children with epilepsy had lower slow wave activity at the start of non-rapid eye movement (NREM) sleep, though similar overnight decline and slow wave activity in the final hour of NREM sleep. Both groups displayed an antero-posterior shift in peak slow wave activity overnight, though individual patients showed persistent local increases at scalp locations matching those of focal interictal discharges. Patients who had seizures during their admission had lower early-night slow wave activity, the group without seizures showing similar activity to controls. We found a positive correlation between full scale IQ and early-night slow wave activity in patients but not controls.

**Interpretation:** Reduced early night slow wave activity in children with focal epilepsies is correlated with lower cognitive ability and more seizures and may reflect a reduction in learning-related synaptic potentiation.

## Introduction

Children with epilepsy display high rates of impairment across neuropsychological domains^1^. The origins of these impairments remain disputed and a range of factors have been implicated, including underlying aetiology, ongoing seizures, and use of antiepileptic drugs^2,3^. More recently, it has been hypothesised that sleep disruption by epileptic activity could play a key mechanistic role^4–6^.

The EEG in deep sleep is dominated by slow waves of high amplitude, generated by widespread cortical neurons alternating in synchrony between depolarized and hyperpolarized states.^7^ Slow wave activity (0.6 - 4.5 Hz) correlates closely with sleep need, building up with time spent awake and dissipating with sleep^8^, thereby providing a quantitative measure of sleep homeostasis. It has been proposed that the decrease in global slow wave activity across the night reflects the process of synaptic renormalisation, which restores cellular functioning and promotes memory consolidation — the ‘synaptic homeostasis hypothesis’. ^9^ There is evidence also that slow wave activity can increase or decrease locally in response to focal stimulation, or intensive training on specific tasks in adults^10,11^ as well as children.^12^ A causal role for slow waves in the consolidation of memory has also been proposed — the ‘active system consolidation hypothesis’ — in which cortical slow oscillations drive and synchronise the replay of neuronal sequences associated with learning during wakefulness, leading to the reactivation and redistribution of temporary, hippocampus-dependent memories to longer-term cortical representation^13^.

Over the course of infancy and childhood, there is evolution of both EEG spectral power and topology^14^. This is particularly marked in the slow wave activity band in sleep, in which we see a ten-fold decrease in power and an occipital to frontal shift in the location of maximal power between preschool age and late adolescence,^15^ in parallel with the acquisition of visual acuity and executive function at those respective ages.^14^

While it is well established that children with epilepsy — particularly those with co-morbid intellectual difficulties — have very high rates of parent-reported sleep disturbance, ^16,17^ the influence of epileptic activity on sleep homeostasis in the developing brain and the relationship of this to cognitive and behavioural measures have thus far not been investigated. Data from adult patients with focal epilepsy suggest that in the mature brain, slow wave activity may be upregulated both globally and locally in response to the burden of epileptic activity.^18^

Here, we investigated sleep homeostasis, as reflected in slow wave activity, in a group of children with focal epilepsy as compared to age-matched controls. We explored the influence of focal epileptic activity on the topological distribution of slow wave activity, and the relationship between sleep homeostasis and cognition. We hypothesised that sleep homeostasis may represent an important mechanism for recovery from the metabolic demands of epileptic activity, and that healthier slow wave activity would correlate with better cognition and behaviour, as well as a lower propensity to seizures.

## Methods

### 1. Participants

Participants were 22 children with drug-resistant focal epilepsy of structural (or presumed structural) aetiology and 21 healthy, age-matched control subjects. Patients were recruited prospectively from the EEG video telemetry unit at Great Ormond Street Hospital for Children (GOSH), London, United Kingdom. Inclusion criteria were: attendance at mainstream school (as a proxy for sufficient cognitive ability to complete the experimental tasks), and planned hospital admission lasting a minimum of four days. Those with a prior diagnosis of primary sleep disorders were excluded. Age and gender-matched healthy control subjects were recruited prospectively by advertisement directed at staff working at Young Epilepsy, a charity that supports children and young people with epilepsy in the United Kingdom. Controls attended the EEG department of the Young Epilepsy campus in Lingfield, United Kingdom to be set up for a single night ambulatory sleep study. All participants were instructed not to nap during the day.

The majority of patients (15/22 patients) were not sleep-deprived. One patient was partially sleep deprived two days before the study night followed by one recovery night (patient 2) while 6 patients (patients 1, 3, 7, 15, 21 and 22) underwent partial sleep deprivation on the study night. This entailed staying up 2 to 3 hours past their habitual bedtime, with recovery sleep truncated at 06:00 the following morning. Four patients (patients 7, 9, 12 and 14) had seizures during the study night.

The study was approved by the National Research Ethics Service and registered with the Joint Research and Development Office of UCL Institute of Child Health and Great Ormond Street Hospital for Children. Written informed consent was obtained from a parent of each participant.

### 2. EEG acquisition and processing

EEG/polysomnography acquisition, visual sleep scoring and the visual quantification and marking of seizures and interictal discharges have been described in previous work.^19^ Controls underwent one night of EEG recording, while in patients, the data acquired across the “sleep” condition of the memory task (as described in section 3.2 below) was used for further analysis. EEG data were recorded with the Xltek Trex (Natus, USA) system, using eight EEG electrodes (F3, F4, C3, C4, O1, O2, A1, A2) positioned according to the International 10-20 system in controls, and 27 EEG electrodes (Fz, Cz, Pz, Fp1, Fp2, F3, F4, F7, F8, F9, F10, C3, C4, C5, C6, T5, T6, T7, T8, T9, T10, P3, P4, P9, P10, O1, O2) positioned according to the International 10-10 system in patients, with the reference electrode at CPz. EEG signal sampling occurred at 512 or 1024 Hz in patients and 256 or 512 Hz in controls.

EEG data were exported to Matlab (version R2015b; The MathWorks Inc., Natick, MA) for processing and analysis. Data were digitally bandpass filtered between 0.3Hz and 40Hz then downsampled to 128 Hz. We performed spectral analysis offline using a fast Fourier transform routine. Power spectra of consecutive 30-second epochs (Hanning window, averages of six 5-second epochs) were computed, resulting in a frequency resolution of 0.2 Hz. The lowest two frequency bins (0.2 and 0.4 Hz) were not used for further analysis due to their sensitivity to low frequency EEG artefacts.^20^ Slow wave activity was defined as power in the 0.6-4.4 Hz range. For each channel, epochs containing artefacts were rejected by visual inspection and semi-automatically, whenever power in the slow wave activity and 20-30 Hz bands exceeded a threshold based on a moving average determined over fifteen 30-second epochs.^21^

All analyses were performed on average-referenced EEG. ^18,22^ To describe overnight changes in slow wave activity, we performed analyses on clean epochs from the first hour versus the last hour of NREM sleep. ^23^ Patient 21, who was awoken less than 3 hours after falling asleep, was excluded from all correlation analyses. ‘Slow wave activity decline’ was defined as the difference between slow wave activity in epochs from the first hour and epochs from the last hour. ‘Normalised slow wave activity decline’ was defined as the ‘slow wave activity decline’ divided by slow wave activity in the first hour of NREM sleep. For group level comparisons between patients and controls, mean slow wave activity was calculated using the 8-electrode average-referenced montage in both groups.

Individual patient topoplots were examined independently by three authors (TB, SC and ME) to determine the overlap between peak slow wave activity and the interictal focus. The interictal focus was obtained from the clinical EEG report. Final decision required consensus, and any disagreements were resolved through discussion.

### 3. Neuropsychological testing and behavioural data

#### 3.1 Full scale IQ

Control subjects underwent IQ assessment using the two-subtest version of the Wechsler Abbreviated Scale of Intelligence (WASI; Pearson, USA). Individual IQ scores on the Wechsler Intelligence Scale for Children version 4 (WISC-IV; Pearson, USA) were available for 16/22 patients from recent neuropsychological testing performed as part of standard pre-operative assessment.

#### 3.2 Memory consolidation

Participants completed memory tasks designed to assess the contribution of sleep to verbal and visuospatial memory consolidation in children, as described in previous work^19^. Briefly, the verbal task required the children to learn a list of semantically related word pairs, with memory tested using a cued recall procedure. The visuospatial task was a two-dimensional object location task, similar to the card game “pairs”. Parallel versions of the tasks were used for “sleep” and “wake” conditions, in which recall was tested after similar length delays either overnight or in the daytime. The order of the conditions was assigned at random and was balanced across the participants. Age-group versions of the tasks ensured comparable difficulty.

#### 3.3 Questionnaires

Questionnaires in relation to each participant’s sleep habits, behaviour and memory were completed by a parent.

##### 3.3.1 Children’s Sleep Habits Questionnaire

The Children’s Sleep Habits Questionnaire ^24^ (CSHQ) is a retrospective parent-report sleep screening instrument designed for and validated on children aged 4 to 10 years. This was applied to all participants, as a validated questionnaire covering the whole study age range was unavailable at the time. Parents are asked to rate the frequency of various sleep behaviours as they would occur in a typical week. 35 items cover the domains of bedtime resistance, sleep onset delay, sleep duration, sleep anxiety, night wakings, parasomnias, sleep-disordered breathing, and daytime sleepiness, and yield a ‘total sleep disturbance score’. A cut-off of 41 is recommended for sleep clinic referral.

##### 3.3.2 Strengths and Difficulties Questionnaire

The Strengths and Difficulties Questionnaire ^25^ (SDQ) is a well-established behavioural screening questionnaire for children aged 3 to 16 years. Twenty-five items cover the domains of emotional distress, behavioural difficulties, hyperactivity and concentration difficulties, getting along with other children and kind and helpful behaviour. Scores in the first 4 domains are added to give a ‘total difficulties score’, for which 0-13 is considered normal and 14-16 borderline.

##### 3.3.3 Everyday Memory Questionnaire

The Everyday Memory Questionnaire ^26^ (EMQ) was originally developed for adults with traumatic brain injury, but has been modified for and validated in children.^27^ Domains covered by the 34 items include memory for faces, places, words, actions and learning new things.

### 4. Statistical analysis

Statistical analyses were performed in R ^28^ and figures were produced using the R package ggplot2 ^29^.

Group differences in demographic data, full scale IQ (FSIQ) and sleep parameters were examined using independent samples t-tests for continuous variables and chi-square tests for categorical variables.

Repeated-measures analyses of variance (ANOVA) with group (patient or control) as the between subjects factor and time (first or last hour of NREM sleep) as the within subjects factor was used to investigate the overnight change in slow wave activity in the patient versus control groups. Normality checks and Levene’s test were carried out and the assumptions were met. Where there were statistically significant interactions, Tukey’s HSD post hoc tests were carried out. An independent samples t-test was used to compare normalised slow wave activity decline in the patient and control groups.

Relationships between slow wave activity characteristics, age, clinical characteristics, memory task performance, neuropsychological functioning and behaviour were examined with Pearson’s r correlation coefficient. A Bonferroni correction was used for multiple comparisons.

Independent sample t-tests were used to evaluate the effect of seizures on slow wave activity.

## Results

Participant demographics and patient clinical characteristics are summarised in Tables 1a and 1b.

**Table 1a.**
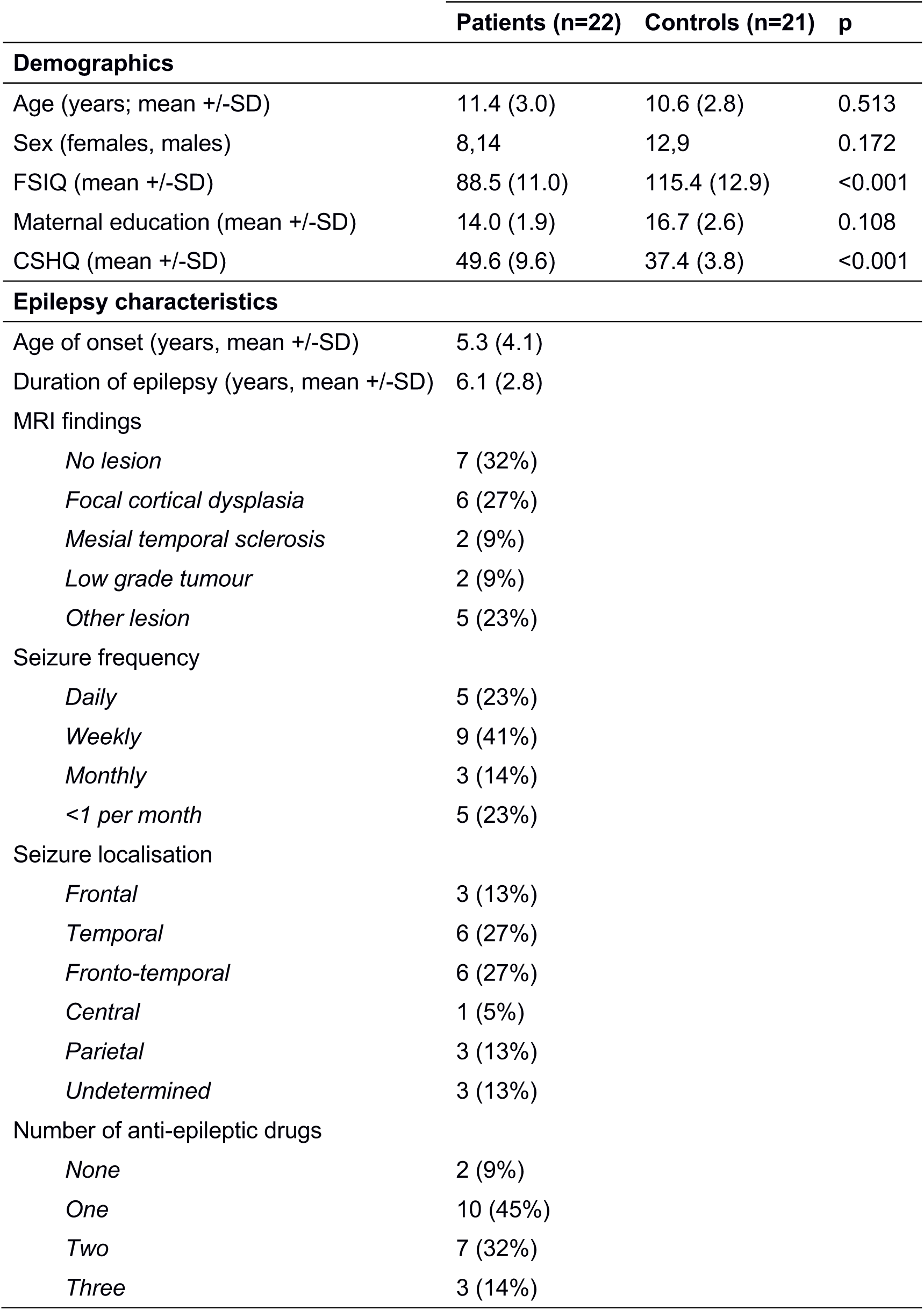
Participant demographics and patient clinical characteristics.

|  | Patients (n=22) | Controls (n=21) | p |
| --- | --- | --- | --- |
| **Demographics** |  |  |  |
| Age (years; mean +/-SD) | 11.4 (3.0) | 10.6 (2.8) | 0.513 |
| Sex (females, males) | 8,14 | 12,9 | 0.172 |
| FSIQ (mean +/-SD) | 88.5 (11.0) | 115.4 (12.9) | <0.001 |
| Maternal education (mean +/-SD) | 14.0 (1.9) | 16.7 (2.6) | 0.108 |
| CSHQ (mean +/-SD) | 49.6 (9.6) | 37.4 (3.8) | <0.001 |
| **Epilepsy characteristics** |  |  |  |
| Age of onset (years, mean +/-SD) | 5.3 (4.1) |  |  |
| Duration of epilepsy (years, mean +/-SD) | 6.1 (2.8) |  |  |
| MRI findings |  |  |  |
| *No lesion* | 7 (32%) |  |  |
| *Focal cortical dysplasia* | 6 (27%) |  |  |
| *Mesial temporal sclerosis* | 2 (9%) |  |  |
| *Low grade tumour* | 2 (9%) |  |  |
| *Other lesion* | 5 (23%) |  |  |
| Seizure frequency |  |  |  |
| *Daily* | 5 (23%) |  |  |
| *Weekly* | 9 (41%) |  |  |
| *Monthly* | 3 (14%) |  |  |
| *<1 per month* | 5 (23%) |  |  |
| Seizure localisation |  |  |  |
| *Frontal* | 3 (13%) |  |  |
| *Temporal* | 6 (27%) |  |  |
| *Fronto-temporal* | 6 (27%) |  |  |
| *Central* | 1 (5%) |  |  |
| *Parietal* | 3 (13%) |  |  |
| *Undetermined* | 3 (13%) |  |  |
| Number of anti-epileptic drugs |  |  |  |
| *None* | 2 (9%) |  |  |
| *One* | 10 (45%) |  |  |
| *Two* | 7 (32%) |  |  |
| *Three* | 3 (14%) |  |  |

**Table 1b.** Patient information.

| Patient ID | Age (years) | Sex | Age of onset (years) | Duration of epilepsy (years) | MRI findings | Seizure frequency | Seizure localisation | Medications |
| --- | --- | --- | --- | --- | --- | --- | --- | --- |
| Patient 1 | 6.6 | Female | 3.0 | 4.6 | Focal cortical dysplasia | Monthly | Fronto-temporal | Carbamazepine |
| Patient 2* | 6.9 | Female | 4.8 | 3.9 | Other lesion | <1 per month | Temporal | Carbamazepine, clobazam, sodium valproate |
| Patient 3 | 7.0 | Male | 6.0 | 6.1 | Focal cortical dysplasia | <1 per month | Frontal | Levetiracetam, sodium valproate |
| Patient 4 | 7.8 | Male | 13.5 | 5.8 | Other lesion | <1 per month | Undetermined | Clobazam, levetiracetam, topiramate |
| Patient 5 | 8.2 | Male | 2.0 | 6.7 | No lesion | <1 per month | Fronto-temporal | Sodium valproate, levetiracetam |
| Patient 6 | 8.8 | Female | 3.0 | 7.3 | Mesial temporal sclerosis | Weekly | Temporal | Levetiracetam, sodium valproate |
| Patient 7 | 9.2 | Female | 6.5 | 7.2 | No lesion | Weekly | Undetermined | Clobazam |
| Patient 8 | 9.8 | Female | 2.0 | 5.0 | Other lesion | Weekly | Parietal | Levetiracetam |
| Patient 9 | 10.4 | Male | 13.0 | 7.4 | No lesion | Daily | Frontal | Carbamazepine |
| Patient 10 | 11.1 | Male | 1.5 | 9.1 | No lesion | Weekly | Fronto-temporal | Carbamazepine, topiramate |
| Patient 11 | 11.3 | Male | 2.0 | 3.3 | No lesion | <1 per month | Undetermined | Levetiracetam, sodium valproate |
| Patient 12* | 11.6 | Male | 12.0 | 5.1 | No lesion | Daily | Fronto-temporal | No medication |
| Patient 13 | 12.6 | Male | 4.0 | 10.6 | Low grade tumor | Weekly | Temporal | Levetiracetam |
| Patient 14 | 12.6 | Male | 8.0 | 8.6 | Other lesion | Daily | Fronto-temporal | Lamotrigine |
| Patient 15 | 12.8 | Male | 2.0 | 4.8 | No lesion | Daily | Frontal | Lacosamide |
| Patient 16* | 14.5 | Female | 2.5 | 3.5 | Focal cortical dysplasia | Daily | Central | Carbamazepine |
| Patient 17 | 14.6 | Female | 2.0 | 1.6 | Low grade tumor | Weekly | Parietal | Lamotrigine, levetiracetam |
| Patient 18 | 14.8 | Female | 1.5 | 2.8 | Other lesion | Weekly | Parietal | No medication |
| Patient 19 | 14.8 | Male | 8.0 | 6.8 | Focal cortical dysplasia | Weekly | Fronto-temporal | Levetiracetam, oxcarbazepine, sodium valproate |
| Patient 20 | 14.9 | Male | 8.0 | 12.4 | Mesial temporal sclerosis | Weekly | Temporal | Oxcarbazepine |
| Patient 21* | 15.3 | Male | 11.0 | 1.8 | Focal cortical dysplasia | Monthly | Temporal | Lamotrigine, sodium valproate |
| Patient 22 | 15.5 | Male | 0.9 | 9.5 | Focal cortical dysplasia | Monthly | Temporal | Lamotrigine |

\* Denotes patients who contributed data to statistical analyses involving demographic data and memory performance, but who were excluded from analyses involving SWA power and topography
Patient 2 - non-contemporaneous sleep and memory data
Patient 12 - visual sleep scoring not possible due to high IED load
Patient 16 - no sleep EEG
Patient 21 - sleep restricted; awoken after less than 2 hours' sleep

### 1. Sleep parameters

Compared to controls, patients had a shorter total sleep time (*M* = 566, *SD* = 51 minutes vs *M* = 483, *SD* = 104 minutes, *t*(30) = 3.35, *p* = 0.002) though sleep efficiency was similar between the two groups (*M* = 93.4, *SD* = 7.03% vs *M* = 91.4, *SD* = 5.72%, *t*(34) = 0.97, *p* = 0.34). Controls and patients spent similar amounts of time in NREM sleep (*M* = 383, *SD* = 35 minutes vs *M* = 356, *SD* = 73 minutes, *t*(35) = 1.56, *p* = 0.13) and showed similar REM latency (*M* = 141, *SD* = 61 minutes vs *M* = 171, *SD* = 81 minutes, *t*(38) *= −1.38, p* = 0.18), though patients spent a smaller proportion of sleep time in REM (*M* = 25.7, *SD* = 4.73% vs *M* = 18.4, *SD* = 7.2%, *t*(36) = 3.89, *p* < 0.001). The visual inspection of the EEG record revealed normal morphology of sleep features, including slow waves and spindles.

### 2. Overnight changes in slow wave activity

We found an interaction between the effects of time and group on slow wave activity, *F*(1, 72) = 4.737, *p* = 0.033. There was a fall in slow wave activity between the first and last hours of NREM sleep in both the controls (*t*(18) = 6.18, *p* < 0.001) and patients (*t*(27) = 4.27, *p* = 0.0015). Controls showed more slow wave activity in the first hour of NREM sleep than patients (*t*(31) = 2.41, *p* = 0.021), but no significant difference in slow wave activity in the last hour (*t*(28) = 0.31, *p* = 0.76), and there was no significant difference in normalised slow wave activity decline (t(25) = −1.61, *p* = 0.12; see Fig 1B).

**Figure 1.**
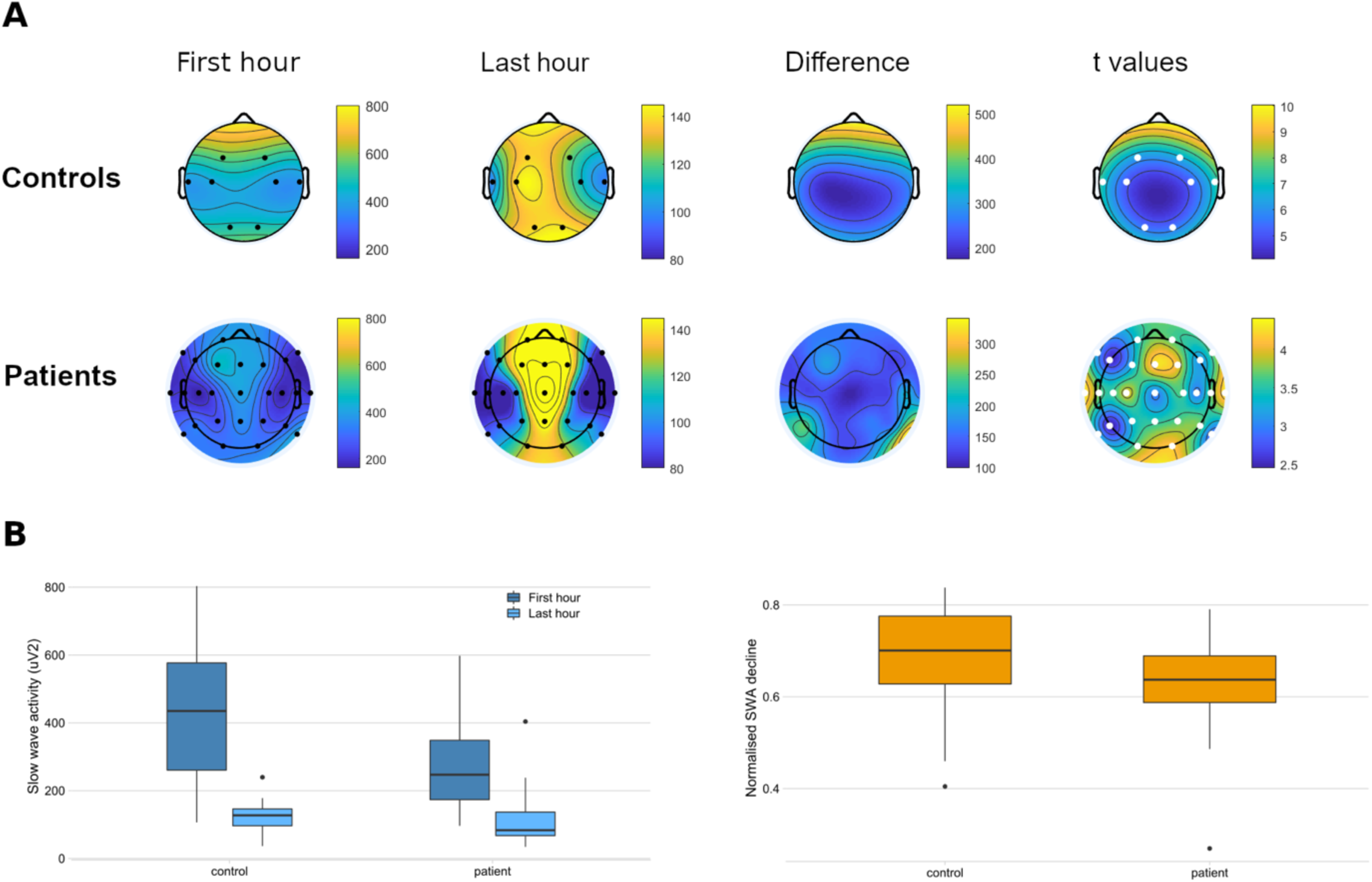
Slow wave activity across the night. (A) Group level slow wave activity topographies in controls (above) and patients (below). There was a significant fall in slow wave activity across the night at all electrodes, and an anteroposterior shift in the slow wave activity peak across the night in both groups. (B) Mean slow wave activity across electrodes in the 8-electrode montage. Left: Early (first hour of NREM sleep) and late (last hour of NREM sleep) night slow wave activity. Controls showed higher slow wave activity early in the night (*p* = 0.021) but slow wave activity was similar by the last hour (*p* = 0.76). Right: Normalised slow wave activity decline (calculated as the difference between early and late night slow wave activity, divided by early night slow wave activity) was not significantly different between patients and controls (*p* = 0.12).

### 3. Topography of slow wave activity

Slow wave activity in the first hour of NREM sleep was highest in the anterior scalp regions in both the control and patient groups. By the last hour of sleep, the peak had shifted to the central or posterior regions. A significant decline in slow wave activity across the night was seen at all electrodes (Fig 1A).

### 4. Correlation between slow wave activity and age

In the control group, we found a decline in slow wave activity with age. This was true of the first and the last hours of NREM sleep [*r*(16) = −0.68, *p* = 0.002; *r*(16) = −0.61, *p* = 0.007]. The magnitude of the decline in slow wave activity across the night also showed an inverse correlation with age [*r*(16) = −0.62, *p* = 0.02]. However, we found no correlation between slow wave activity and age in the patient group (Fig 2).

**Figure 2.**
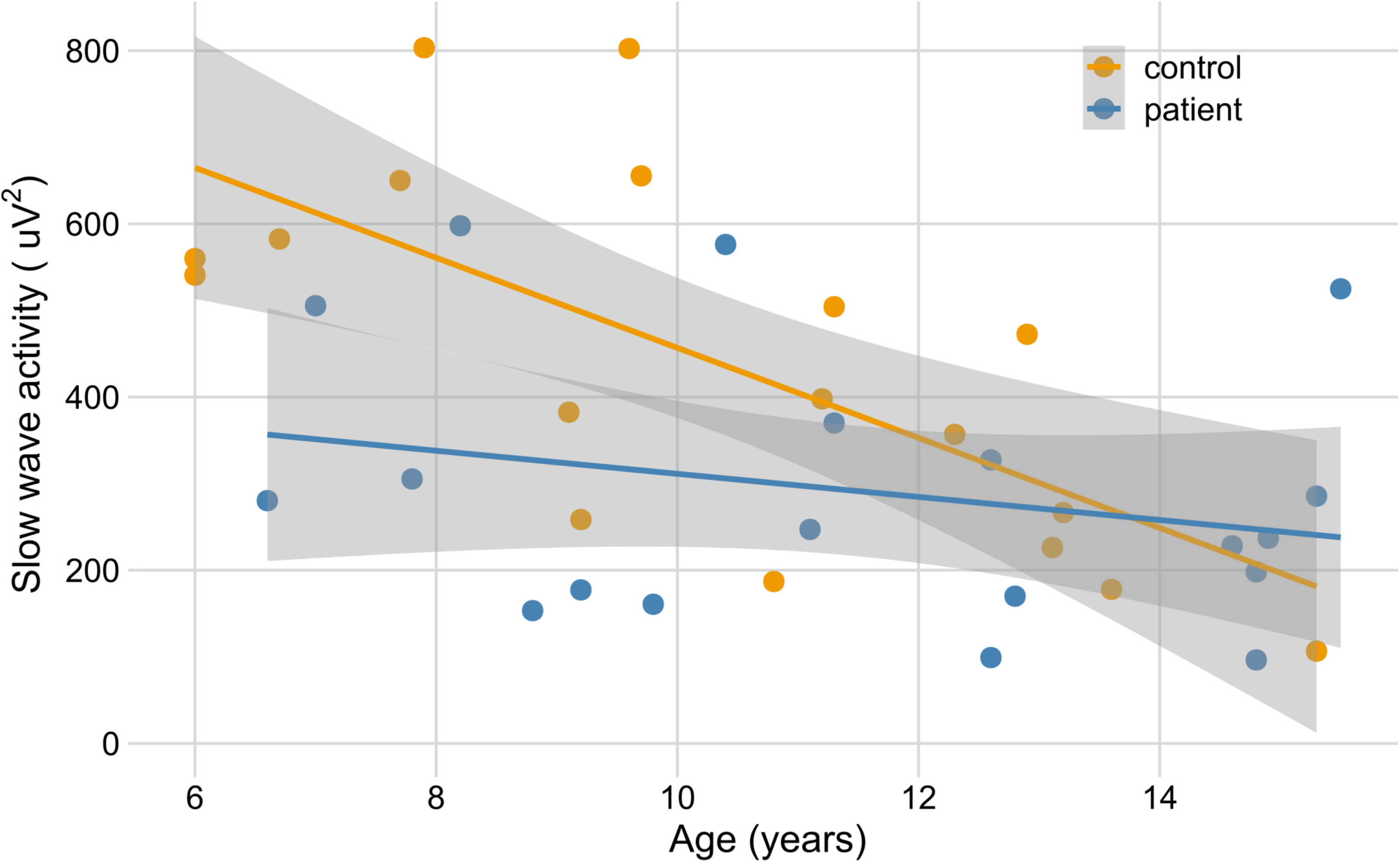
Decline in slow wave activity with age. Early night slow wave activity decreased with age in healthy control subjects [*r*(16)= −0.68, *p* = 0.002], but this correlation did not reach significance in the patients [r(17)=-0.26, *p* = 0.29].

### 5. Effect of epileptic activity on slow wave activity

In the first hour of NREM sleep, the location of maximal slow wave activity coincided with the foci of interictal discharges in 13 of 19 patients. In the last hour of NREM sleep, this was true in 15 of 19 patients (Fig 3).

**Figure 3.**
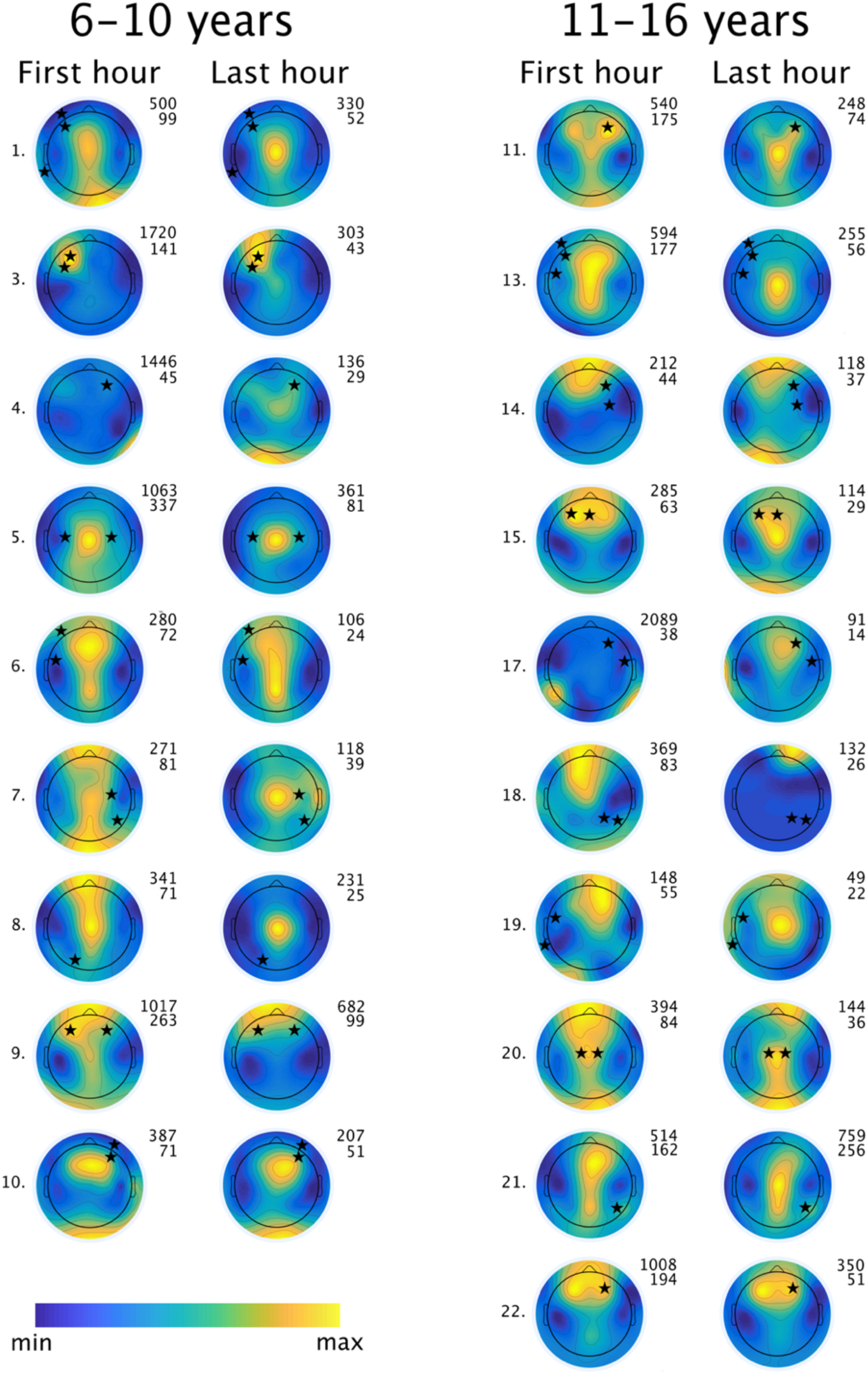
Changes in local slow wave activity overnight. Individual topographies for all patients with available EEG data, arranged in order of increasing age. Black stars indicate foci of interictal discharges as stated on the clinical EEG report. In the majority of patients (15/19), peak slow wave activity persists at or moves towards the interictal foci in the last hour of NREM sleep. Patients 6, 7, 9, 10, 13, 14, 15, 17, 19 and 20 had seizures during their admission, while the others did not. Patients 1, 3, 7, 15, 21 and 22 underwent sleep restriction.

Patients who had seizures during their hospital admission showed less slow wave activity in the first hour of NREM sleep than controls (*M* = 192.9, *SD* = 74.1 µV^2^ vs *M* = 440.6, *SD* = 212.6 µV^2^, *t*(26) = −3.07, *p* = 0.004), while those who did not have seizures during the admission showed similar slow wave activity to controls (*M* = 358.6, *SD* = 152.4, *t*(21) = −1.15, *p* = 0.26). Only 2 of the 10 patients with seizures had seizures prior to the study night. The difference between patients with and without seizures reached statistical significance providing the outlier was removed (*t*(16) = −1.89, *p* = 0.076; *t*(11) = −2.91, *p* = 0.013 without Patient 9; see Fig 4).

**Figure 4.**
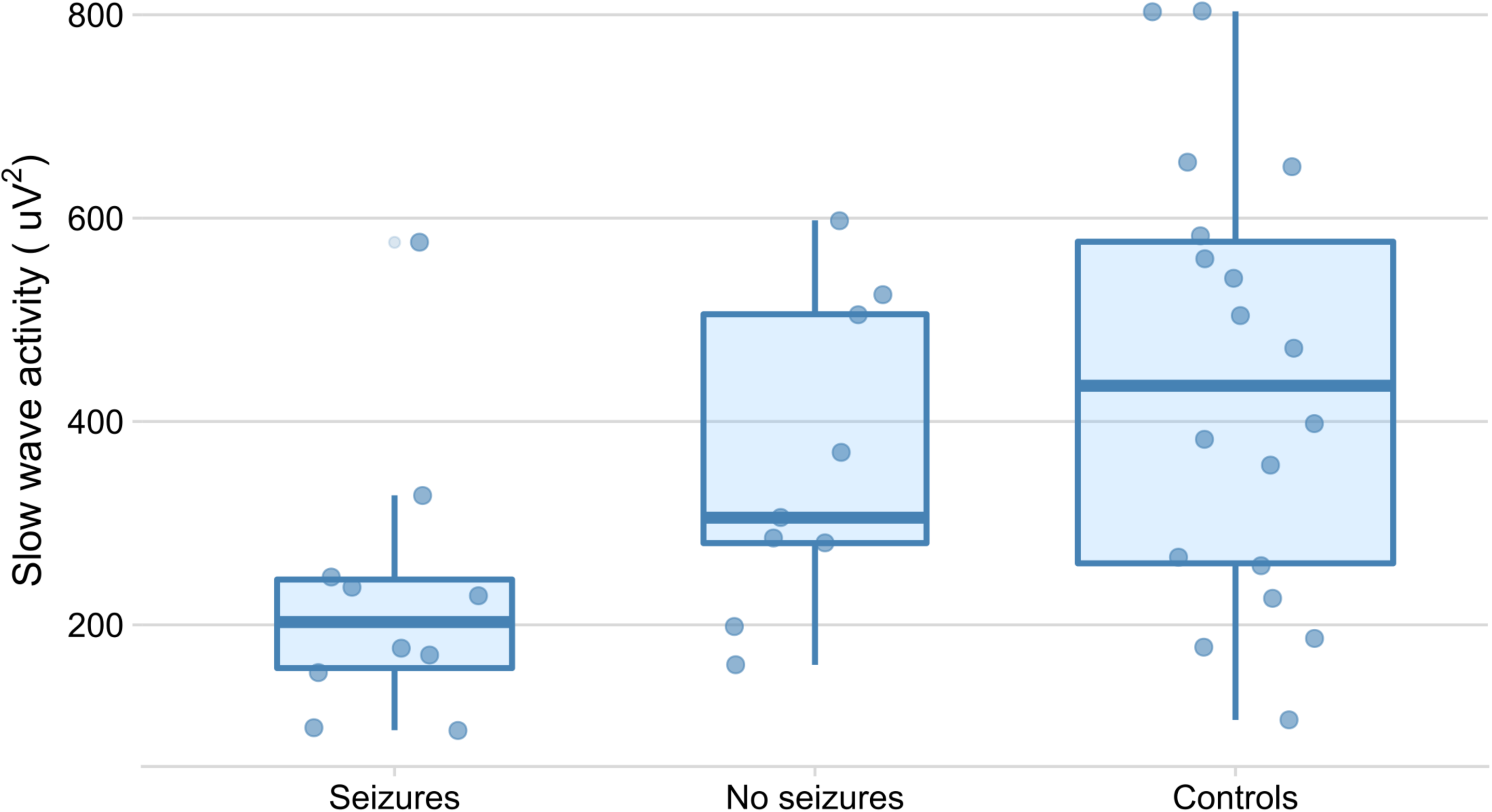
Slow wave activity and seizure propensity. Patients who had seizures during their hospital admission showed less slow wave activity in the first hour of NREM sleep than controls (*p* = 0.004), while those who did not have seizures during the admission showed slow wave activity not significantly different from controls (*p* = 0.26). Box widths are proportional to group size and jitter plots show data for individual participants.

Nine of the 10 patients with seizures during admission showed a coincidence of the late night slow wave activity peak with the interictal discharge focus, while this was true of only 5 of the 9 patients without seizures (Fig 3). The patients who underwent sleep restriction (patients 1, 3, 7, 15, 21 and 22) did not show higher early night slow wave activity than those who were not sleep restricted (Fig 3).

### 6. Relationship of slow wave activity to cognition

We found no significant correlation between slow wave activity or overnight slow wave activity decline and memory task performance on overnight recall, or change in recall between the ‘wake’ and ‘sleep’ conditions in either memory task across the total participant sample or within each group.

Higher slow wave activity in the first hour of NREM sleep was correlated with greater FSIQ in the patient group [*r*(14) = 0.57, *p* = 0.02; adjusted *p* = 0.04], but not in the controls [*r*(15) = −0.11, *p* = 0.68] (Fig 5). There was no correlation of FSIQ with overnight slow wave activity decline.

**Figure 5.**
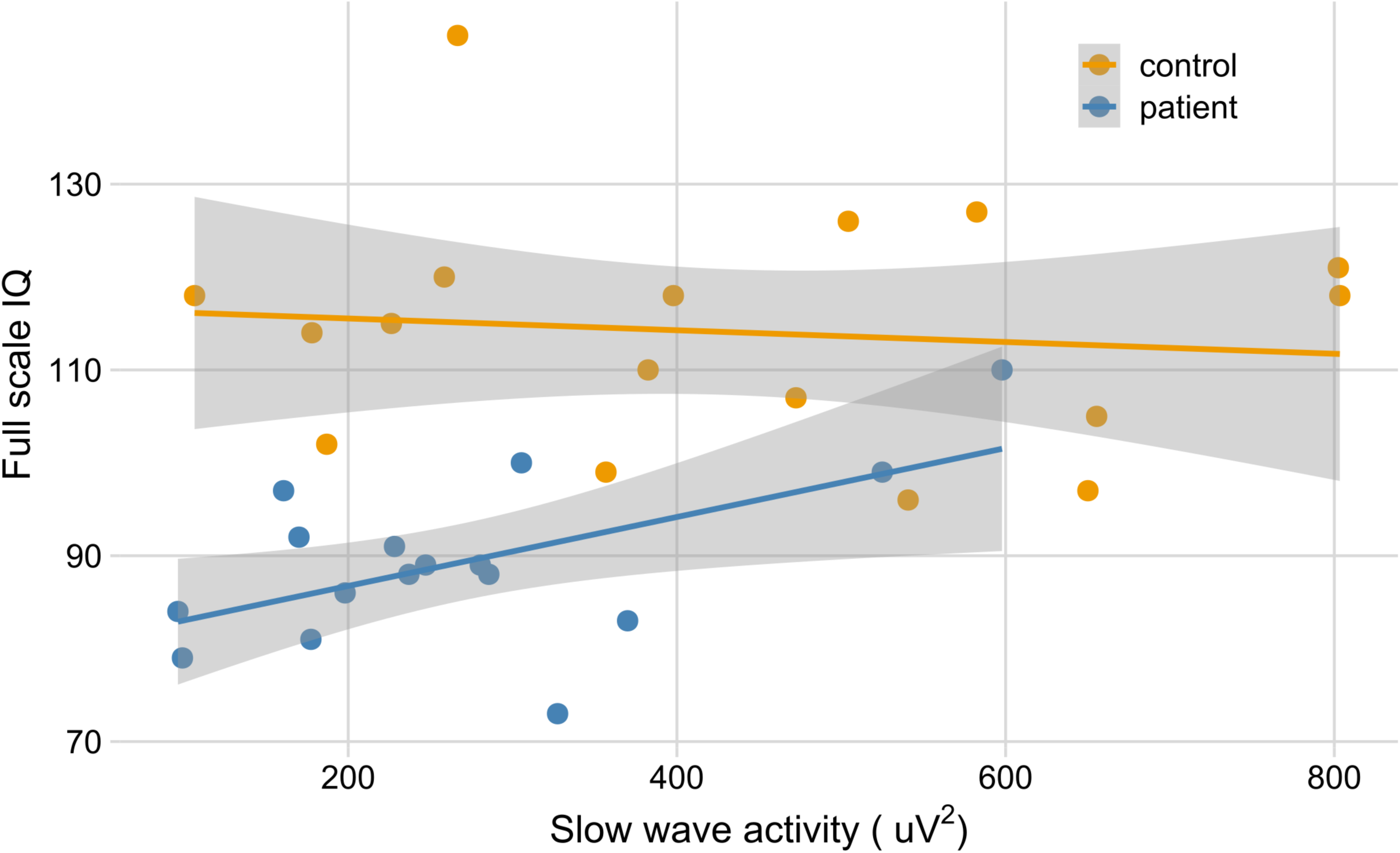
Correlation between FSIQ and early night slow wave activity. FSIQ correlated with early night slow wave activity in patients [*r*(14) = 0.57, *p* = 0.02; adjusted *p* = 0.04] but not in controls [*r*(15) = −0.11, *p* = 0.68].

### 7. Relationship of slow wave activity to behavioural questionnaire scores

We found no correlation between slow wave activity and behavioural questionnaire scores.

## Discussion

We have performed the first study in children relating the influence of epileptic activity on sleep homeostasis to cognitive and behavioural measures. Compared to age-matched healthy controls, children with focal epilepsy showed less slow wave activity at the start of the night, though similar decline in slow wave activity overnight. Within the patient group, lower early night slow wave activity was associated with a higher likelihood of subsequent seizures and lower FSIQ.

### Slow wave activity build-up and decline

Slow wave activity at the start of the night reflects sleep need,^8^ and has been shown to build up with prolonged wakefulness, both in adults ^30^ and, more recently in school-aged children. ^31^ While slow wave activity in the control group was consistent with published data from healthy children ^31–33^, the patient group showed lower early-night slow wave activity across all scalp regions. Slow waves had normal morphology, and slow wave activity topology in the patients at group level was age-appropriate both at the start ^14^ and at the end ^31^ of the night.

While the hospital confinement of the patients restricted their physical activity levels, the magnitude of the deficit in early-night slow wave activity seen here was far greater than that of the increase recorded as a consequence of intense exercise.^34^ Possible confounding factors such as pathological slowing or the effects of partial sleep deprivation ^20,31^ would have driven up the slow wave activity. One explanation could be that the physiological synaptic strengthening in response to environmental stimuli which underpins learning during wakefulness ^9^ may be suppressed. This would be associated with less spatial and metabolic demand with the result of lower slow wave activity after falling asleep. Alternatively, the ability to generate slow waves may be impaired or immature in patients compared to age-matched controls.^20^

Slow wave activity shows an exponential decline across the night, ^20,30^ though we have here made a linear approximation (“normalised decline”) using measurements from the first and last hours of NREM sleep. ^23,35^ We found that children with focal epilepsy showed similar normalised slow wave activity decline to controls over the course of the night. This suggests that synaptic downscaling is successfully achieved. ^9^ According to the synaptic homeostasis hypothesis, this downscaling of synapses underpins the benefit of sleep on memory consolidation, providing a mechanistic explanation for our previous finding that sleep benefits memory consolidation in this patient cohort.^19^

### Effect of epilepsy on maturational trends in slow wave activity

We demonstrated a striking decline in early-night slow wave activity with age across our cohort of healthy controls, replicating the findings of Kurth et al. ^15^ and Feinberg et al. ^36^ However we did not find a correlation of early-night slow wave activity with age in the patient group, suggesting an increased variability in the homeostatic response to synaptic strengthening during wakefulness. This heterogeneity may explain the divergence of our findings from those of Boly et al., ^18^ who demonstrated increased slow wave activity in a group of adult patients with focal epilepsies compared to controls.

Those in whom we recorded no seizures during their admission showed similar early-night slow wave activity to controls, while those experiencing seizures during admission had lower early-night slow wave activity. We might conclude that high seizure propensity is associated with impaired learning-related potentiation of synapses (and potentially impaired cognition) and, as a consequence, early night slow wave activity is lower. Interestingly, most of the seizures occurred after the study night, raising the possibility that lower early night slow wave activity could be a marker of increased seizure propensity.

Topological plots of slow wave activity at the individual level showed that in some patients, peak slow wave activity coincided with and persisted at the foci of interictal discharges across the night, obscuring any age-related trends. ^15^ Data from healthy adults show that slow wave activity increases locally in response to intense training in a task involving specific cortical regions,^10^ and can be interpreted as a response to local use-dependent strengthening of synapses. In congruence with our findings, a local increase in slow wave activity coinciding with the seizure focus has been demonstrated in some adults with focal epilepsy.^18^ Though the interictal discharge counts in this cohort were extremely low, with a median of just 1 discharge per 2.48 minutes, ^19^ the fact that patients with seizures were more likely to show a persistence of the slow wave activity peak at the foci of interictal discharges suggests that this may represent a local strengthening of pathological networks. ^4,35^

### Cognitive correlates

We found a correlation between early-night slow wave activity and FSIQ in the patient group, but not in the controls. This suggests that suppressed synaptic potentiation during wakefulness leading to lower early night slow wave activity could be a sign of reduced or inefficient learning, contributing to reduced crystallised intelligence in the longer term. Conversely, patients with lower IQ may have less capacity for learning during wakefulness, resulting in less synaptic potentiation, thus perpetuating the intellectual deficit. The absence of this relationship in the control group may reflect a ceiling effect of FSIQ.

We found no correlation between early-night slow wave activity or normalised slow wave activity decline and performance in the memory tasks, though we have previously reported the correlation between sleep benefit on the verbal task and slow wave sleep duration.^19^ Wilhelm et al. ^37^ demonstrated a correlation between slow wave activity across NREM sleep over the whole night and performance on a motor sequence task the following day in healthy adults. Both slow wave sleep duration and whole-night slow wave activity are dependent on the amount of sleep time occupied with cortical slow oscillations, thus their positive correlation with memory task performance is consistent with the active system consolidation hypothesis.^13^

### Behavioural correlates

There was no correlation between slow wave activity and behavioural questionnaire scores, including the Children’s Sleep Habits Questionnaire, though parents of patients reported more behavioural and sleep difficulties in their children than the parents of controls.^19^ This suggests that behaviour does not have a simple relationship with slow wave activity dynamics.

### Limitations

The present study has several limitations. The patient group was heterogeneous in their baseline clinical characteristics, and having been admitted to hospital to capture ictal EEG, underwent interventions including antiepileptic drug reduction and sleep restriction. It is thus difficult to comment on the differences in sleep architecture compared to controls, although it is notable that sleep efficiency and REM latency were similar. It is also unlikely that medication effects would have led to a systematic bias of the results since patients were on different medications with an even spread between those which increase and those which decrease SWS. ^38^ Our analysis of slow wave activity topology in controls was limited due to the 8-electrode montage, however we have replicated the findings of others in terms of slow wave activity and age-related trends in healthy children. More accurate results regarding focal changes in slow wave activity topology in the patients could be sought using high density EEG.

### Conclusion

Our findings provide novel insight into the possible mechanisms by which sleep may influence cognition in children with epilepsy. Slow wave activity build-up, presumably reflecting the learning related increase in synaptic strength over waking hours is reduced in patients, interestingly, more so in the ones with higher seizure propensity and lower cognitive abilities. On the other hand, the overnight slow wave activity decline, reflecting synaptic recovery and thought to be responsible for the beneficial effect of sleep on memory consolidation is preserved in this patient group, who show few epileptic discharges during sleep. Studies of the awake EEG record during learning as well as within-patient comparisons of sleep records where the ability to generate slow waves is stressed may yet further our understanding.

## Data Availability

Data not available due to ethical restrictions.

## Acknowledgements

We are grateful to Cleo Chevalier-Riffard, Hannah Scrivener, Holly Sayer and Ralph Smith for their assistance with data collection and preprocessing, to staff and participants at Young Epilepsy for facilitating the control arm of the study. SC is funded by a University of London Chadburn Lectureship. This research was funded by Action Medical Research and the Reta Lila Howard Foundation and supported by the National Institute for Health Research Biomedical Research Centre at Great Ormond Street Hospital for Children NHS Foundation Trust and University College London. This work was supported by the Starr International Foundation and the Anna Müller Grocholski-Foundation to BKB. TB is supported by the Great Ormond Street Hospital Children’s Charity.

## Author contributions

SC and BKB conceived and designed the study. ME, SC, BKB and TB analysed the data. ME and SC wrote the manuscript and produced the figures. All authors have edited and approved the final draft of the manuscript.

